# COVID-19 Compromises in the Medical Practice and the Consequential Effect on Endometriosis Patients

**DOI:** 10.1101/2021.05.04.21255000

**Authors:** Shaked Ashkenazi, Ole Linvåg Huseby, Gard Kroken, Adrian Soto-Mota, Marius Pents, Alessandra Loschiavo, Roksana Lewandowska, Grace Tran, Sebastian Kwiatkowski

## Abstract

**Background and purpose:** In response to the ongoing coronavirus disease 2019 (COVID-19) pandemic, self-isolation practices aimed to curb the spread of COVID-19 have severely complicated the medical management of patients suffering from endometriosis and their physical and mental well- being. Endometriosis, the main cause for chronic pelvic pain (CPP), is a highly prevalent disease characterized by the presence of endometrial tissue in locations outside the uterine cavity that affects up to 10% of women in their reproductive age. This study aimed to explore the effects of the global COVID-19 pandemic on patients suffering from endometriosis across multiple countries, and to investigate the different approaches to the medical management of these patients based on their self-reported experiences.

**Methods:** A cross-sectional survey, partially based on validated quality of life questionnaires for endometriosis patients, was initially created in English, which was then reviewed by experts. Through the process of assessing face and content validity, the questionnaire was then translated to fifteen different languages following the WHO recommendations for medical translation. After evaluation, the questionnaire was converted into a web form and distributed across different platforms. An analysis of 2964 responses of participants from 59 countries suffering from self-reported endometriosis was then conducted.

**Results:** The data shows an association between COVID-19 imposed compromises with the reported worsening of the mental state of the participants, as well as with the aggravation of their symptoms. For the 1174 participants who had their medical appointments cancelled, 43.7% (n=513) reported that their symptoms had been aggravated, and 49.3% (n=579) reported that their mental state had worsened. In comparison, of the 1180 participants who kept their appointments, only 29.4% (n=347) stated that their symptoms had been aggravated, and 27.5% (n=325) stated their mental health had worsened. 610 participants did not have medical appointments scheduled, and these participants follow a similar pattern as the participants who kept their appointments, with 29.0% (n=177) reporting aggravation of symptoms and 28.2% (n=172) reporting that their mental state had worsened.

**Conclusions:** These findings suggest that COVID-19 pandemic has had a clinically significant negative effect on the mental and physical well-being of participants suffering from endometriosis based on their self-reported experiences. Thus, they show the importance of further assessment and reevaluation of the current and future management of this condition in medical practices worldwide.

## BACKGROUND

Existing literature demonstrates that the quality of life of women suffering from endometriosis was impaired in a multitude of ways, even before the COVID-19 pandemic (Moradi *et al*., 2014; Arion *et al*., 2020). These include but are not limited to reduced work productivity (de Graaff *et al*., 2013), as well as negative effects on relationships, education, and general well-being (Soliman *et al*., 2017; van Poll *et al*., 2020). Although numerous studies on quality of life of patients suffering from endometriosis have been undertaken, many of them have a relatively small sample size (de Graaff *et al*., 2013; González-Echevarría *et al*., 2019; Corte *et al*., 2020).

The rapid spread of coronavirus disease 2019 (COVID-19) around the globe has triggered dramatic and often transformational effects on routine health care practices (Birkmeyer *et al*., 2020). COVID-19-related policies and recommendations have further reduced the availability of caregivers and compromised healthcare for patients suffering from a variety of conditions (Wallis *et al*., 2020). In particular, the Obstetrics and Gynecology practice has been compromised across multiple countries (“Quality of life and quality of society during COVID-19 | Eurofound,” n.d.).

Many medical centers have temporarily ceased offering surgical management for endometriosis, which is a crucial part of the management of the condition (Parasar *et al*., 2017), and appointments for outpatient settings are currently being postponed or cancelled (OECD/European Union, 2020). These factors negatively impact the standard of care for these patients.

Additionally, the International Society of Ultrasound in Obstetrics and Gynecology has recommended postponing ultrasound evaluation of non-acute pelvic pain (Bourne *et al*., 2020).

Furthermore, endometriosis patients have reported their concerns with seeking medical help out of fear of getting infected with SARS-CoV-2 in medical centers (Leonardi *et al*., 2020a). Consequently, the quality of life of endometriosis patients has been drastically impaired by pain, subfertility, frustration about disease recurrence, and uncertainty regarding the therapeutic options available to them (Ammar *et al*., 2020; Pfefferbaum and North, 2020). These restrictions were reported to put endometriosis patients at risk of negative psychological effects additional to those inflicted by mandated self-isolation (Gordon and Balsom, 2020).

This study aimed to explore the effect of the global coronavirus disease 2019 pandemic on patients suffering from endometriosis across multiple countries, and to investigate the different approaches to the medical management of these patients based on their self-reported experiences.

## METHODS

The methodological design of this study involved two phases: phase 1 was qualitative, and phase 2 involved a cross-sectional survey.

A computerized search of PMC-US National Library of Medicine, BMC Women Health, and Health Affairs resources was performed to identify registered articles about endometriosis and Obstetrics and Gynecology management published before and during the current global pandemic, as well as registered articles regarding COVID-19 and the healthcare system management. The search was conducted using the following terms: “Endometriosis and quality of life”; endometriosis and COVID-19”; and “Healthcare and COVID-19”.

The literature review included comparative studies, qualitative studies, clinical trials, controlled and randomized controlled trials, and multicenter studies. Several articles were selected on the basis of inclusion criteria and cross-references checked. Upon finalization of the initial English survey, translations of the survey to fifteen languages were initiated, including: Arabic, Farsi, Finnish, French, German, Greek, Hebrew, Italian, Norwegian, Polish, Portuguese, Russian, Spanish, Swedish and Turkish.

Translations were aimed at the conceptual equivalent of relevant phrases and words, as recommended by the World Health Organization criteria (World Health Organization, 2017), yet avoided “word- for- word” or literal translation. It aimed for all 3 phases of forward translation, expert panel, and back translation for every language.

### Questionnaire structure

The questionnaire comprised four distinct sections. The first section collected basic information about the respondents, including their age, nationality, and country of residence during the global pandemic. This information has not violated their anonymity; rather, it has enabled a categorization of the responses based on these details for a later evaluation.

The second section of the questionnaire was based on a review of EHP-30 (Khong *et al*., 2010), a validated tool designed to measure the health-related quality of life (HRQoL) in women with endometriosis (Bourdel *et al*., 2019; Moradi *et al*., 2019; Weeks, 2020). This section inquired about general patient- and disease-specific characteristics in order to determine the current specific condition that the respondent is diagnosed with in addition to when they were diagnosed, the effect of endometriosis on their life, and how it might limit their activity. This section also included questions regarding the respondent’s current treatments, including fertility treatments.

The third section of the questionnaire investigated the effects of the global pandemic on respondents, incorporating “yes/no” questions. An option of “this is irrelevant for me” was added in correspondence to the specific question. This section investigated whether the respondent had experienced any cancelation/ postponement of appointments that were initially scheduled for the diagnosis, treatment, or both of their endometriosis and in vitro fertilization (IVF) appointments of a variety of kinds.

The fourth section of the questionnaire was measured on a numerical rating scale. The degree to which the respondents agreed with the statement given in each question was scored on a scale ranging from 1 to 5, with 1 representing “strongly agree” and 5 representing “strongly disagree.” Each question incorporated statements regarding the effects of the pandemic on a respondent’s decision to seek medical help concerning their endometriosis condition, aggravation of symptoms due to the current global situation, changes in the respondent’s mental state, as well as statements concerning the medical management of their disease during the pandemic.

The questionnaire was converted into an online self-administered survey, which was distributed among the participants via email, social media platforms, and academic circles through multi-center collaborations. Data was collected from these online self-administered surveys, and subsequently interpreted for further analysis.

### Ethical approval

The bioethics committee of the Pomeranian Medical University in Szczecin provided an exemption from an ethical consent-case number: KB-0012/34/03/2021/Z. Additionally, this study was also granted an ethical approval from the Turkish Ministry of Health: 2021-01-13T17_02_26, Başvuru Formu için tıklayınız// KONU No: KAEK/2021.01.27.

### Data analysis

Chi-square tests for independence were carried out for the analysis of the two variables: suspension of health services, and the patients mental and physical well-being.

## RESULTS

Out of 3024 participants from 59 countries who submitted the questionnaire between November 2020 and January 2021, 2964 (98.01%) provided information that enabled the proper analysis of the results. Table 1 shows the demographic and clinical characteristics of the participants. As described, the mean age of the participants is 33.2 (SD: +/- 7.5) and the distribution between the stages is as follows-stage 1: 4,8% (n=142); stage 2: 9% (n=267); stage 3: 14,7% (n=435) and stage 4: 30,7% (n=910). 40,8% (n=1210) of participants stated that they are not currently diagnosed with a specific stage of the disease.

**TABLE 1.**
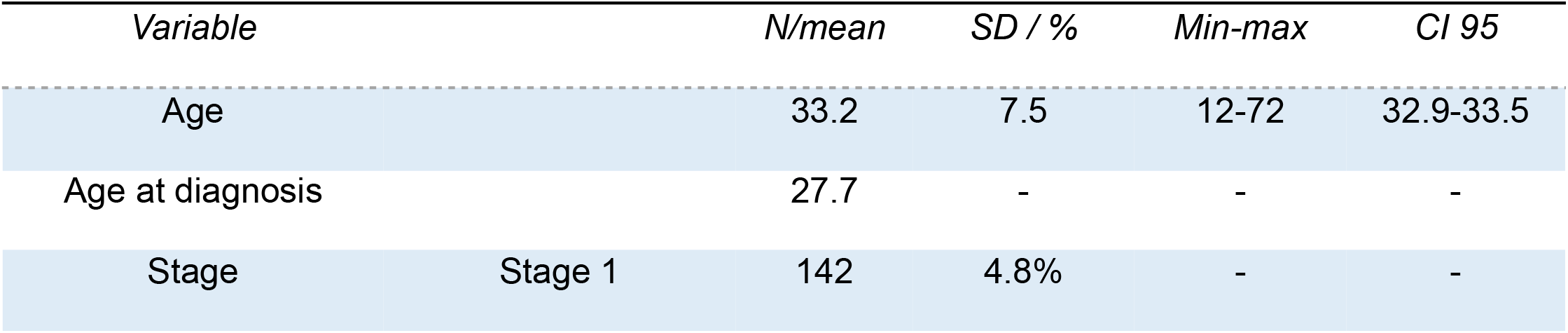
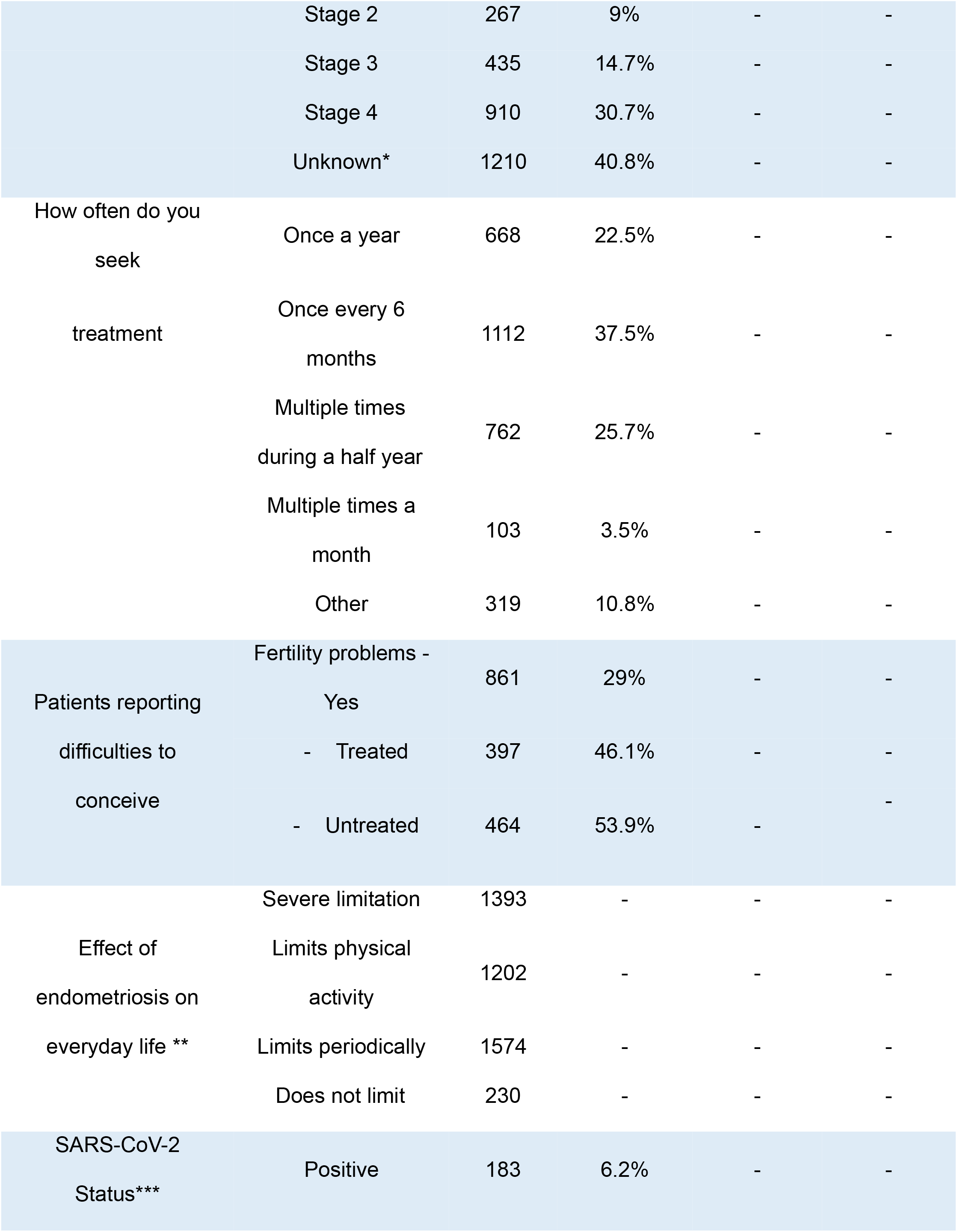
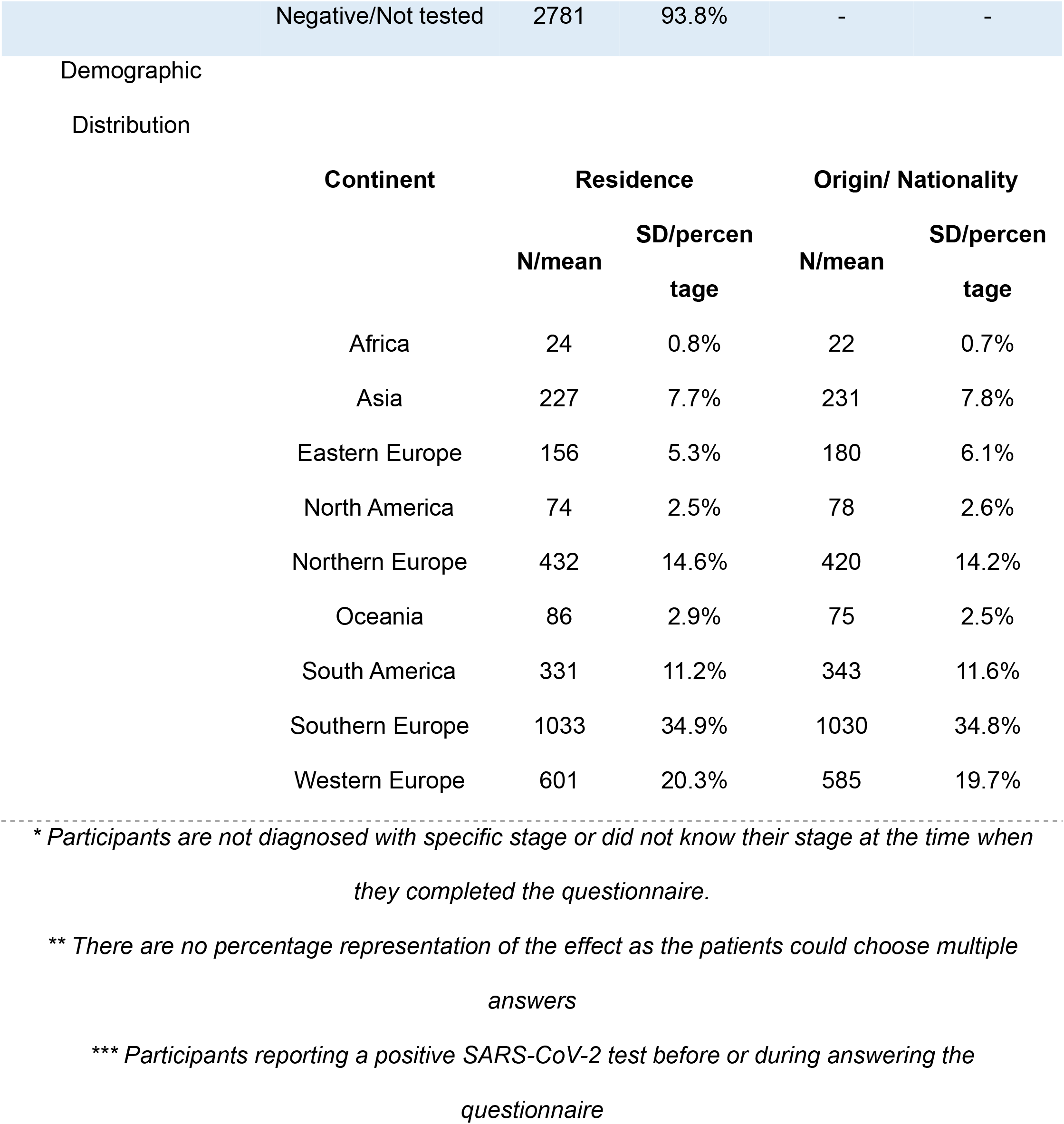

Furthermore, this table outlines the general and transformational effects that endometriosis imposes on the everyday life of participants. Only 230 participants have stated that their condition has no significant effect or no effect at all on their everyday activities, while 1393 participants have stated that they experience a severe compromise in their routine activities. This table also summarizes the frequency with which the participants seek medical attention concerning their endometriosis, the fertility status of the participants, as well as the distribution of participants who were previously or currently diagnosed with SARS-CoV-2.

Table 2 shows the reported mental health changes that the participants experienced during the COVID-19 pandemic at the time of completing the questionnaire. Figure 1 outlines the demographic distribution of the participants, related to their reported worsening of mental and physical well-being.

**TABLE 2.**
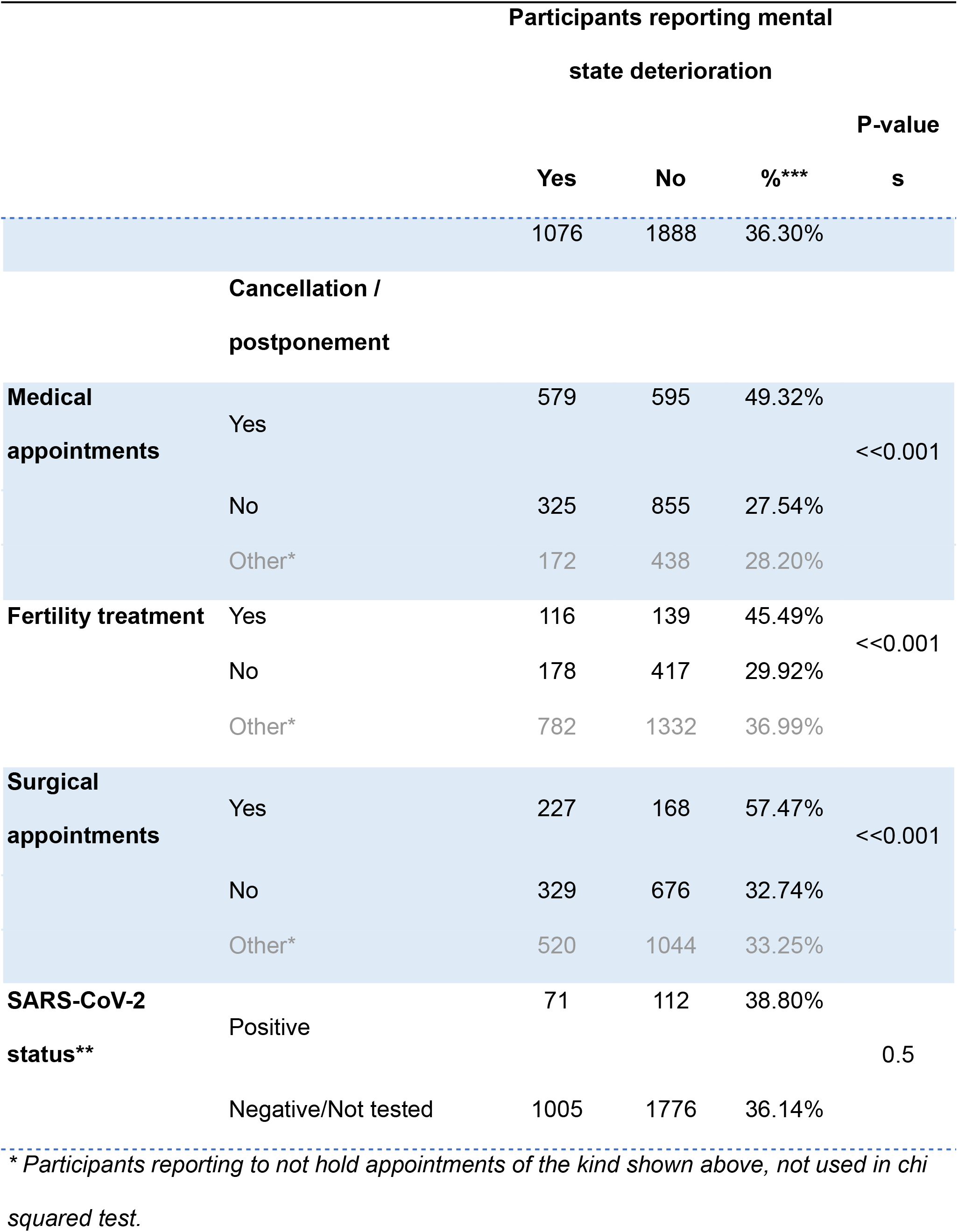
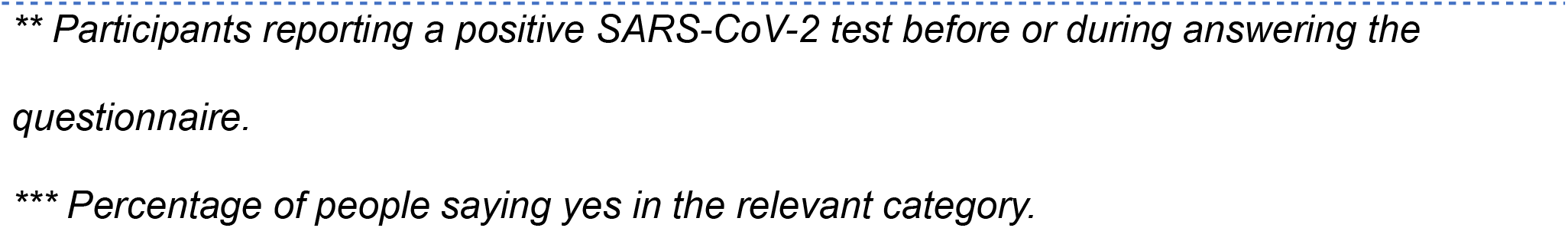

**Figure 1.**
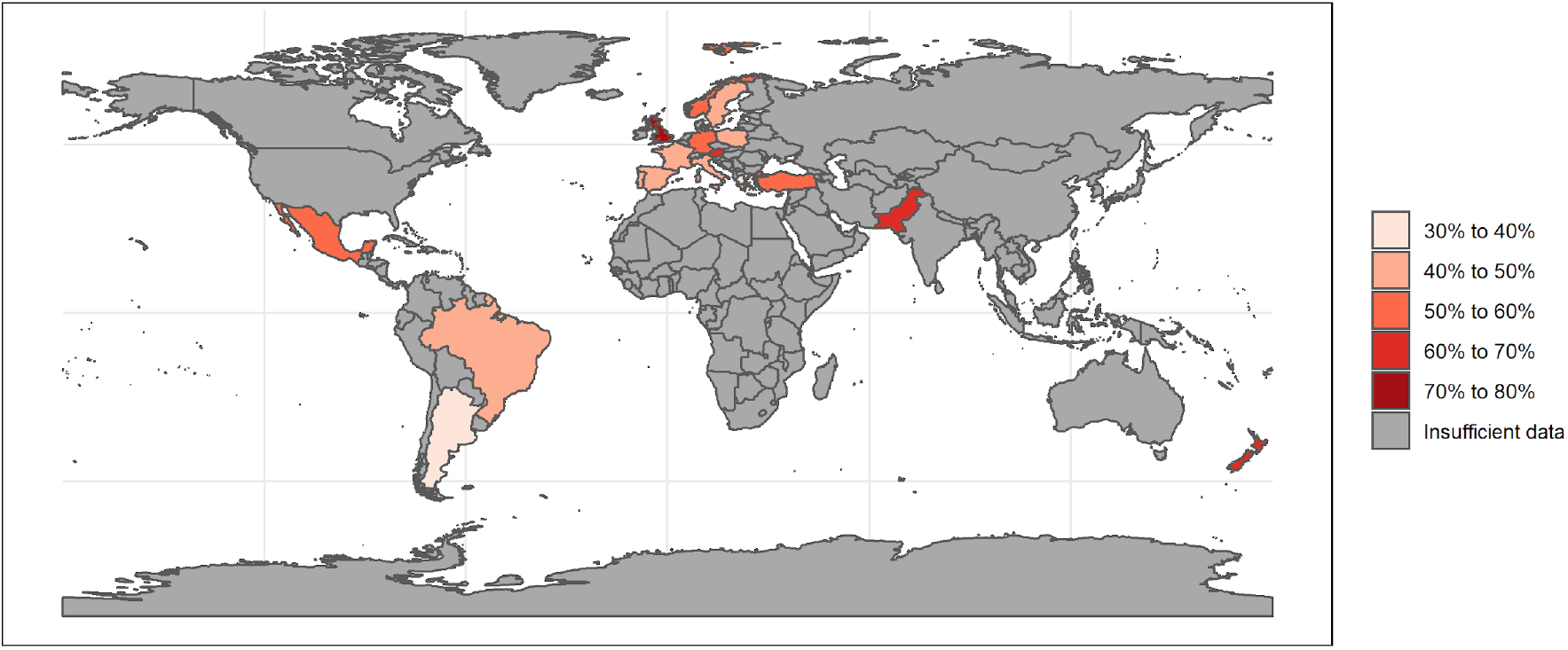

36.3% of the participants reported that their mental health had worsened during the pandemic. 1174 participants reported some kind of cancellation to medical appointments, and 49.3% of them stated that their mental health had deteriorated. In comparison, of the 1180 participants who did not experience cancellations and the 610 who did not have any scheduled appointments, 27.5% and 28.2% reported worse mental health, respectively. Moreover, the table identifies the number of participants who reported that their scheduled fertility treatment and/or surgical appointments were postponed or cancelled. It also shows that among the participants reporting worsening of mental health, 38.8 % (n=71) have tested positive for SARS-CoV-2, and 36.1% (n=1005) have tested negative or were not tested at all.

Table 3 shows that 35.0% of participants feel their symptoms have been aggravated during the COVID-19 pandemic. For the 1174 participants who had their medical appointments cancelled, 43.7% reported that their symptoms had been aggravated. In comparison, from the 1180 participants who kept their appointments and the 610 that did not report to have any medical appointments scheduled, 29.4% and 29.0% stated that their symptoms had been aggravated, respectively. Table 3 also identifies the number of participants that reported that their scheduled fertility treatments and/or their surgical appointments were postponed or canceled. Moreover, from the participants reporting symptomatic aggravation, 36.0% have tested positive for SARS-CoV-2, and 34.9% have tested negative or were not tested at all.

**TABLE 3.**
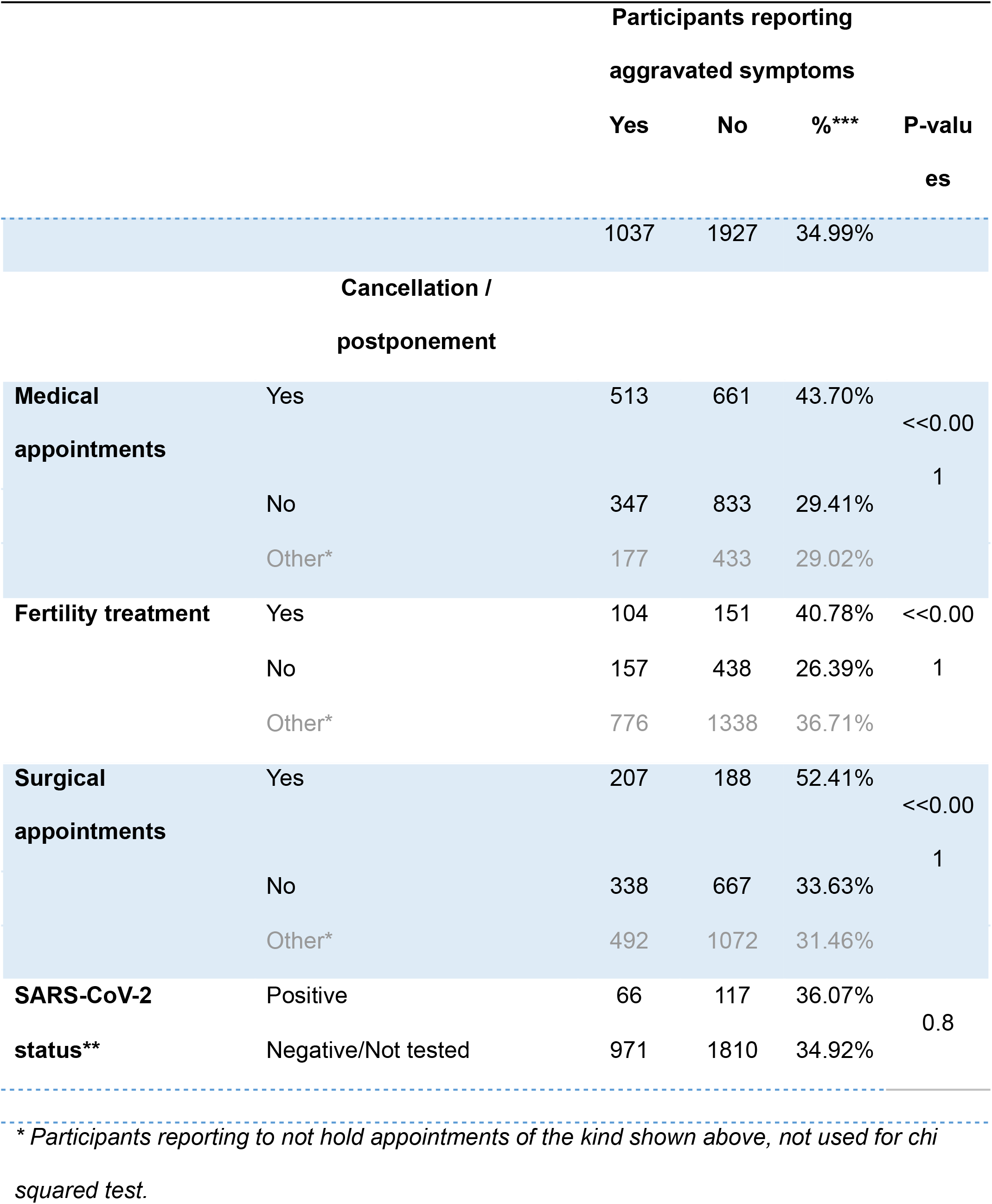
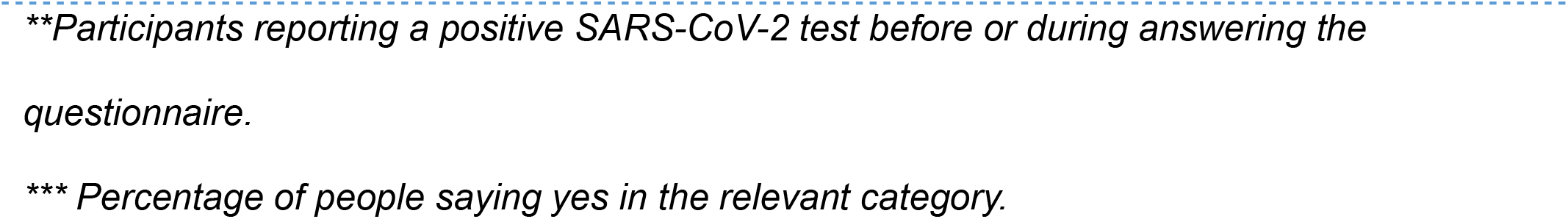

Tables 4a and 4b summarize the self-reported impact of the COVID-19 pandemic on medical healthcare and overall well-being. Most of the respondents (79%, n=2358) had at least one healthcare appointment scheduled during the pandemic. Almost half (49.9%, n=1176) of them reported at least one cancellation and almost 30% of the scheduled surgical and fertility treatments were cancelled (28.2%, n=396 and 29.9%, n=255 respectively). Additionally, almost half of the participants (48,8%, n=961) reported they would have sought emergency gynecological attention but refrained from doing so because of their fears concerning arriving at a medical institution at the time of the global pandemic.

**TABLE 4.**

|  | Yes | No | Does not<br>apply | % cancelled<br>appointments |
| --- | --- | --- | --- | --- |
| Cancellations of any kind | 1176<br>(39.6%) | 1182<br>(39.8<br>%) | 611<br>(20.5%) | 49.9 |
| Cancelled surgeries | 396<br>(13.3%) | 1007<br>(33.9<br>%) | 1566<br>(52.72%) | 28.2 |
| Cancelled fertility treatments | 255<br>(8.5%) | 597<br>(20.1<br>%) | 2117<br>(71.3%) | 29.9 |
| Refrained from seeking<br>emergency gynaecological<br>attention | 961<br>(32.3%) | 1010<br>(34.0<br>%) | 998<br>(33.6%) | 48.8 |

**B)**
|  | Strongly agree | Agree | Neither | Disagree | Strongly disagree | % Agree or strongly agree |
| --- | --- | --- | --- | --- | --- | --- |
| Refrained from seeking any gynecological attention | 560<br>(18.9%) | 441<br>(14.9%) | 674<br>(22.7%) | 431<br>(14.5%) | 863<br>(29.1%) | 33.7 |
| Would seek more help without the pandemic | 695<br>(23.4%) | 495<br>(16.7%) | 471<br>(15.9%) | 347<br>(11.7%) | 961<br>(32.4%) | 40.1 |
| Symptoms aggravated during the pandemic | 601<br>(20.2%) | 438<br>(14.8%) | 650<br>(21.9%) | 383<br>(12.9%) | 897<br>(30.2%) | 35.0 |
| Mental state worsened during the pandemic | 568<br>(19.1%) | 509<br>(17.1%) | 536<br>(18.1%) | 428<br>(14.4%) | 928<br>(31.3%) | 36.3 |
| Their condition would have been managed better without the pandemic | 714<br>(24.0%) | 450<br>(15.2%) | 523<br>(17.6%) | 389<br>(13.3%) | 893<br>(30.1%) | 39.2 |

More than a third of the respondents reported that their symptoms or mental well-being deteriorated during the pandemic (35%, n=1039 and 36%, n=1077 respectively) and 39% (n=1164) of them believed that their condition would have been managed better if the COVID-19 pandemic had not occurred. Conversely, 43% (n=1282) respondents asserted that the ways in which they manage their endometriosis have not particularly changed because of or during the COVID-19 pandemic. 18% (n=523) of the participants reported neither an impairment of their endometriosis management, nor lack of change.

## DISCUSSION

This study represents an open window to a problem that has been evident even before the pandemic, in which compromising resources to treat and diagnose endometriosis significantly affects the overall quality of life of those who suffer from it (Leonardi *et al*., 2020b). The pandemic has amplified existing compromises on the general resilience of health care systems worldwide, especially as they relate to the management of space, human, and material resources (OECD/European Union, 2020).

Drawing on data from 2964 participants from 59 countries, who are diverse in ethnicity, nationality, and socio-economic status provided the opportunity to present a well-established estimation accounting for the above-mentioned factors. As expected, our data indicate that general absence of care directly impacts quality of life for patients suffering from endometriosis.

Our study detected important alterations in respondents’ mental and physical well-being, and almost 50% reported a decline in either or both during the COVID-19 pandemic. Our findings suggest that this reported decline in physical and mental well-being can be attributed to the cancellation / postponement of medical appointments, including surgical and fertility treatments. This is supported by other studies which have reported considerable negative impacts on women’s mental health and quality of life while they await fertility treatment during the COVID-19 pandemic (Gordon and Balsom, 2020).

Similarly, 28% have had their scheduled surgical appointments delayed, which can postpone both proper treatment and diagnosis. Reports are conflicting about the relevance of these delays in healthcare patients (Unger and Laufer, 2011; Hudelist *et al*., 2012). Nonetheless, several studies have reported a delay in diagnosis of 7 to 12 years in women with endometriosis (Hadfield *et al*., 1996; Husby *et al*., 2003; Ballard *et al*., 2006; Staal *et al*., 2016). Thus, it is fair to assume that even further delay in both the diagnosis and treatment can be expected during the COVID-19 pandemic. Further follow-up is needed to learn the true impact that this will have.

Most procedures and appointments in endometriosis healthcare are elective. However, the fact that almost 40% of respondents believe that their condition would have been better managed were it not for the COVID-19 pandemic indisputably deserves attention. Importantly, more than a third of the participants reported physical or mental harm attributable to the pandemic on their healthcare, and the consequences of these detriments are yet unknown. Long term follow-up studies will also be needed to assess this.

Finally, it is concerning that almost half of the participants refrained from seeking emergency gynecological attention. It remains possible that future phases and implementations of social restrictions will be required. Since all healthcare systems should be prepared to face future high demand challenges, it is necessary to design and implement strategies to allow all non-COVID-19 emergencies to be properly managed.

Lacking direct and easily quantifiable outcomes, it will be particularly difficult to estimate the consequences of the COVID-19 pandemic on people living with non-lethal, highly prevalent chronic diseases such as migraine, fibromyalgia, and endometriosis.

Despite the inherent differences between these illnesses, it is likely that at least some of the repercussions for endometriosis patients that we documented are reflected in other diseases with the aforementioned characteristics. Insight from this study should prove useful for updating endometriosis clinical management guidelines all around the world, and for improving the resilience of healthcare systems against future high-demand challenges.

### Limitations of the study

Due to a low number of SARS-CoV-2 positive respondents, any test of association would be underpowered, and we are therefore unable to say whether there is a significant connection or not.

With an international questionnaire, arising issues of cultural differences and subjective answers are likely inevitable. In most cases, the research team ensured that at least two people who spoke the target language were translating the survey from English to the target language, but could not always ensure that two translators whose mother tongue was English were also both fluent in the target language.

Further limitations with a multiple-choice questionnaire are that participants can allude to different meanings when selecting the same answer. This problem increases when trying to reach an international sample of people. Furthermore, the questionnaire was anonymous, and we have no confirmation of whether the participants are indeed real and whether they answered honestly, although there is little reason to suspect otherwise, given that there was no incentive to take this questionnaire.

Distribution of the survey online, through various platforms and with the help of national and international endometriosis organizations, resulted in varying levels of success, and in some countries, we did not manage to release the questionnaire at all.

Europe and South America are more represented than other areas, with around 90% of the respondents residing in these continents.

### Author’s notes

The intention of this paper is therefore not to focus on the differences between countries, but on the general rather than specific effects of absence of care. Considering the study’s statistical qualities, the findings are unlikely coincidental. While the global COVID-19 pandemic is ongoing, the present study’s findings are not limited to COVID-19 alone but enable us to understand the consequences of general absence of care in many forms and to eventually conclude how to better manage chronic diseases in the future, and in relationship to endometriosis.

### Future directions

Further research is needed to assess the true impact and long-term consequences of the COVID-19 pandemic for patients living with endometriosis. For now, simpler measurements can be implemented to mitigate the detrimental effects that limited health care has had on the reported health of the participants. Telemedicine with video consultations shows promise for some patients (Grimes *et al*., 2020). This cannot replace necessary face-to-face consultations like surgical procedures, but can perhaps help patients that have suboptimal treatment, as they can be followed up digitally.

## CONCLUSION

There are multiple components affecting the quality of life of women suffering from endometriosis. Our study reveals a clear correlation between the deterioration of the reported physical and mental state, and impaired medical care for patients suffering from endometriosis during the COVID-19 pandemic. The largest difference in reported well-being was found among patients who were supposed to undergo surgical procedures but had their appointments cancelled or postponed due to the pandemic.

## Supporting information

Acknowledgement

## Data Availability

Data is intended to be shared upon request to the corresponding author if applicable.

