## Supplementary material for "COVID-19 Compromises in the Medical Practice and the Consequential Effect on Endometriosis Patients": Acknowledgement

**Translation:**

**Arabic-** Sahl Challar, M.D.* (2023) Pomeranian Medical University in Szczecin Mohammed Aljaff, M.D.* (2023) Pomeranian Medical University in Szczecin, Khalil Gharios, Business Administration, (class of 2020), Coventry University, UK.

**Finnish-** Aurora Henriksson, M.D.* (2023) Pomeranian Medical University in Szczecin, Elena Lekkas, M.D.* (2026) Pomeranian Medical University in Szczecin,

**French-** Mithula Shanmugasuntharam, M.D.* (2023) Pomeranian Medical University in Szczecin, Laura Besenbruch, Actress in Paris.

**German-** Isabella Kuhn, M.D.* (2023) Pomeranian Medical University in Szczecin, Miriam Hehlmann, M. Sc., University of Trier in Germany

**Greek-** Roxani - Magdalini Sidirokastriti, MA (Translation Studies), University of Birmingham U.K, Marina Lemis, Law LLB University of Reading UK, JMW Solicitors LLP, Anna Diakou, Law LLB University of Reading UK, Slaughter and May

**Hebrew -** Shaked Ashkenazi, M.D* (2023) Pomeranian Medical University in Szczecin, Hadas Yeverechyahu, Tel-Aviv University

**Italian-** Alessandra Loschiavo**,** M.D* (2021/2022) Università degli Studi della Campania Luigi Vanvitelli, Marta Lo Presti, UniCredit Bank - Milan

**Norwegian-** Ole Linvåg Huseby, M.D* (2023) Pomeranian Medical University in Szczecin, Daniel Hoseth Nilsen, M.D. (2018) Pomeranian Medical University in Szczecin

**Persian**-Leila Amini, PhD, Nursing and amidwifery School, Iran University of Medical Sciences, Iran, Tahmine Salehi, Tahmine Salehi, B.S., M.S. in Nursing Management, PhD in nursing. Associate Professor

**Polish-** Roksana Lewandowska, M.D.* (2021), Pomeranian Medical University in Szczecin

**Portuguese-** Beatriz Pestana Figueira Santos Faria, M.D.* (2022), Faculdade de Medicina da Universidade de Lisboa

**Russian-** Smolkin Yaakov, M.D. (2017), Pomeranian Medical University in Szczecin, Nikol M. M, Kolin Zolotnitsky

**Spanish-** Pablo Ignacio Soto Mota, Ph.D. Research Scholar, Norwegian School of Economics, Juan Sebastián Garrigues Vega, M.D* (2024), Pomeranian Medical University in Szczecin

**Swedish-** Kwabena Owusu-Mari, M.D.* (2026), Pomeranian Medical University in Szczecin

**Turkish-** Şeyma Taştekin, M.D.* (2022), Trakya University School of Medicine in Turkey

**Cooperating centers-**

1. Department of Woman, Child, and General and Specialized Surgery- University of Campania Luigi Vanvitelli, Napoli, Italy
2. Department of Hygiene and Epidemiology, Pomeranian Medical University in Szczecin
3. Endometriosis New Zealand
4. World Endometriosis Organizations
5. Turkish endometriosis and Adenomyosis Society

We would like to thank important individuals who, in many different ways, made this study possible and helped us through the way.

1. Dr. Joanna Nieznanowska , our dear teacher
2. Deborah Bush, World Endometriosis Organizations principal, Chief Executive Endometriosis New Zealand, and World Endometriosis Society Board Trustee for her tremendous help, support and true belief in our international study. She always found time to consult, guide and refer the research team to the right sources in order to achieve voluble, credible and significant results. We would also like to thank her for her mission- leading the world to a better understanding of endometriosis and her mission to increase the research of endometriosis
3. Miguel Gallego

Organizations:

1. **Endometriose-Vereinigung Deutschland e.V.**
2. **Association EndoFrance**
3. **ENDOmind Franc**/ endofrance
4. **Endometrioseforeningen Norge**
5. **Endometriosis Israel**
6. **Endometriosis New Zealand**
7. **Endometriosföreningen Sverige**
8. Sana Kardar, Biomedical Science (2015), University of London
9. World Endometriosis Organization- the large global cohort reflected in this research demonstrates how those with endometriosis, research teams and clinicians can collaborate to expand knowledge and insights to advance understanding of endometriosis and its serious human impact. The impact of Covid-19 on those with endometriosis has world-wide relevance in all settings
